# Technical feasibility and acceptance of the Remote Monitoring Application in Psychiatry (ReMAP)

**DOI:** 10.1101/2020.11.04.20225730

**Authors:** Daniel Emden, Janik Goltermann, Udo Dannlowski, Tim Hahn, Nils Opel

**Author notes:** Corresponding author: N. Opel. Department of Psychiatry. University of Münster. Albert-Schweitzer-Campus 1. 48149 Münster. Germany. This is to indicate that DE and JG contributed equally to the present work.

## Abstract

**Background:** Over recent years smartphone-based monitoring has been recognized as a useful instrument in psychiatric research. Due to the phasic character of affective symptoms, mobile assessments of passive sensor data as well as active self-reported data via the participants smartphone might represent a cost-efficient and highly useful tool for prospective prediction of mood changes. Despite these promising opportunities, smartphone-based monitoring in psychiatry is still limited to pilot studies often focusing on a single disorder while large-scale, transdiagnostic studies are widely absent.

**Objectives:** The present paper describes the functionality and development of the Remote Monitoring Application in Psychiatry (ReMAP). We aimed to investigate the technical feasibility, and the acceptance of the ReMAP app for the continuous assessment of affective symptoms among different patient groups.

**Methods:** The ReMAP app was distributed among a sample of n=997 composed of healthy control participants and psychiatric patients. Continuous passive sensor data were assessed comprising acceleration, geolocation, as well as walking distance and steps. Further, participants optionally provided standardized self-reports on mood and sleep, as well as voice samples. Technical feasibility and acceptance were assessed based on the amount and frequency of transferred data events, as well as participation duration. Preliminary results are presented while data collection is ongoing.

**Results:** Retention rates of 90.25% for the required minimum study duration of two weeks and 33.09% for one year respectively were achieved. On average, users participated for 150 days. An average of 51.83 passive events per day per participant was collected, with an average rate of 73.50% of days during participation containing passive events. An average of 34.59 active self-report events were transferred per participant, with a considerable range across participants (0-552 events). While clinical and non-clinical participant subgroups did not differ in participation duration, or in quantity or rate of passive or active data transfer, the rate of days with transferred passive data was considerably higher and less heterogeneous in iOS (mean=91.85%, SD=21.25) as compared to Android users (mean=63.04%, SD=35.09).

**Conclusions:** The ReMAP app is technically feasible and generally well accepted and therefore represents a viable complementary tool for the continuous assessment of affective symptoms in large-scale transdiagnostic psychiatric studies. Future studies should account for the observed systematic differences between operating systems.

## Introduction

Increasing efforts have been made to predict individual trajectories of psychiatric disorders, as well as the response to therapeutic interventions in the context of personalized medicine. In affective disorders the previous course of symptoms is one of the major predictors of the future trajectory and its assessment could thus pose a central role for models of personalized prediction. However, the assessment of symptom trajectories is challenging. Retrospective self-reports of symptomatology may be imprecise or biased due to acute affective symptoms (Coyne, Thompson, & Racioppo, 2001). Further, recurrent cross-sectional assessments could result in an incomprehensive picture of the trajectory if the sampling-rate is too low. Recently, continuous prospective assessments using smartphones have been proposed as a possible approach to fill this gap. Data derived from smartphone sensors and smartphone-delivered self-reports could thus complement a multitude of other data entities that have been proposed as promising/potential targets useful for individual prediction (e.g. neuroimaging, genetics, behavioral/clinical data). The abundance of sensors integrated within modern smartphones has resulted in a vast variety of different data modalities of which the utility has been investigated (Onnela & Rauch, 2016; Russell & Gajos, 2020; Torous et al., 2018). In the context of affective disorders, subjective self-reported mood ratings and objective sensor-derived data have been investigated and the predictive utility of sensor data for mood trajectories has been demonstrated in preliminary studies (Dogan, Sander, Wagner, Hegerl, & Kohls, 2017). For example, acceleration and location data have been utilized to differentiate unipolar and bipolar disorder patients (Faurholt-Jepsen et al., 2012, 2015). Further, suicidal phenotype profiles that were longitudinally associated with suicidal behavior have been developed based on frequent smartphone-delivered surveys of suicidal ideation (Kleiman et al., 2018).

While these preliminary studies provide general proof of concept, the exact benefit of the utilization of smartphone-derived data for psychiatric research is still unclear. The value of statistical models for the prediction of future events is often unknown or very limited, resulting in an urgent need for a more systematic investigation of predictive validity (Torous, Choudhury, Barnett, Keshavan, & Kane, 2020). Further, a number of technical and methodological challenges arise within this relatively young field of research (Torous et al., 2020).

As described in a review by Bardram and Matic (2020) various frameworks and apps for digital phenotyping currently exist - each with different advantages and drawbacks. One important aspect with regard to existing software is that most apps have been designed exclusively for one operating system (Android or iOS) thus considerably limiting the large-scale applicability in research samples. Further, existing frameworks rely on a fixed backend and data management infrastructure that might not be compatible with data security policies e.g. in European countries that require data storage on a server based in the EU. This is particularly problematic as the majority of cloud infrastructure providers are based in the US (Bardram & Matic, 2020). Finally, most frameworks do not implicate third-party developer customization and are therefore not flexible to be customized for individual research requirements (Bardram, 2020). The aspects outlined above illustrate the considerations and challenges that investigators of digital phenotyping are currently facing in psychiatry. Data protection guidelines, customizability, and availability for the two most used operating systems iOS and Android enabling large-scale distribution, were the major reasons for the development of a new app system for our study that is introduced in the current paper.

We developed the smartphone app *Remote Monitoring Application in Psychiatry* (ReMAP) for the assessment of affective symptoms that combines a multitude of passive and active data modalities. ReMAP is used as an add-on assessment that complements data entities derived from existing longitudinal deep phenotyping studies. The utilization of the assessed mobile data requires an investigation of 1) the technical feasibility, 2) the acceptance and adherence, 3) the cross-sectional validity, 4) the predictive validity.

In the current study the development and implementation of ReMAP is described and the technical feasibility (1) as well as the acceptance and adherence of the app (2) are investigated. The cross-sectional validity of mobile assessments of affective symptoms via ReMAP (3) has previously been demonstrated (Goltermann et al., 2020), while future studies will investigate the predictive validity of ReMAP based data (4).

## Methods

### The ReMAP App

#### Overview

The ReMAP mobile application has been developed natively for both iOS and Android at the Institute for Translational Psychiatry at the University of Münster, Germany since mid-2018 and has been continuously enhanced since then to adapt to new operating system versions and devices. It is based on Apple ResearchKit, Apple Health and Google Fit, which has sped up the development time and enables comparability with data from other studies, which use identical underlying frameworks.

After entering the individual Subject ID, the app works in background mode and collects data without any active involvement of the participant. In particular, the app collects the number of steps taken by the user and the distance walked from Google Fit or Apple Health. Accelerometer (x, y, z axis) and GPS are additionally monitored.

The continuous collection of these variables over long periods of time provides insight into activity profiles and patterns of everyday activities in near real time, thus delivering data of high ecological validity. At the same time, the passive data collection approach minimizes attrition rates, avoids missing values and is not burdensome for participants.

In addition to passive data collection, participants are regularly asked to complete surveys. These include self-reports about sleep and mood as well as the Beck Depression Inventory (BDI) (Beck, Ward, Mendelson, Mock, & Erbaugh, 1961) and a short one to three minute voice recording about the participant’s well-being during the past week.

#### Participant pool and Recruitment

The ReMAP study was designed as an add-on assessment for ongoing longitudinal observational studies. Thus, the ReMAP data is surveyed in an array of patient and healthy control subsamples and complements further data entities - i.e. structural and functional MRI data, genetics and microbiome data, neuropsychological tests, clinical interviews, and a variety of clinical and personality questionnaires. Collectively, studies that host the ReMAP survey, include patients with affective disorders, psychotic disorders, and anxiety disorders, as well as healthy control participants without any history of psychiatric conditions (see **Supplementary Material** for further details).

Within the context of these ongoing studies a total of n=2952 participants were approached and disclosed about the ReMAP app between December 2018 and September 2020. Interested subjects were extensively briefed about aims, methods (especially type and amount of collected data), details on data security (details on data transfer and storage), and financial compensation. The latter was provided for passive transfer of data for a duration of at least two weeks and one year respectively and not conditional on active data transfer.

Of all participants that were approached, a total of n=997 approved to additionally participate in the ReMAP survey (installations with valid ID) thus yielding an inclusion rate of 33.77%.

Written informed consent of all participating subjects was obtained and each participant was provided an individual subject ID (subject code).

Participants were asked to download the developed smartphone app ReMAP and to start the application. At this time, subjects were asked to confirm again participation in the study and to enter their individual subject ID. Also, participants needed to confirm that ReMAP is granted access to passive data recorded via Apple HealthKit or Google Fit at the time of the initial activation of the app. Research staff was present and able to assist the participants during this process if needed. After entering the individual Subject ID, the app worked in background mode and collected data without any active involvement of the participant and without draining the battery. All acquired data is encrypted on the smartphone and transferred from the participants smartphone to a backend server located within the university network of the University of Münster via internet connection in a pseudonymised and encrypted form on a regular basis. A financial compensation of 10 Euro was granted if a participant transferred passive data for a required minimum time interval of at least two weeks. Participation beyond the two-week interval was optional with another 10 Euro granted if participants transferred data for an entire year. The study was approved by the local IRB as well as by the local data protection officer.

#### Active Data Types

##### Surveys

For the surveys included in ReMAP, we aimed to develop a framework that works offline and fits well into a native app in terms of design and usability. Offline usability was a prerequisite at the beginning of the project, since it cannot be assumed that the respondent has a stable internet connection at all times. To reduce the development and implementation effort of surveys from scratch, we chose to use Apple ResearchKit for iOS and an open source solution called SurveyKit (https://github.com/quickbirdstudios/SurveyKit) for Android, which is inspired by ResearchKit. This facilitated the implementation because the two frameworks use highly similar interfaces. Merely, the notification system had to be reimplemented for both platforms.

The surveys are managed via the ReMAP backend and defined there as configuration files. For example, if you want to integrate a new survey into your study, you only have to extend the configuration file and it will be loaded at the next app start. In the survey definition all surveys and answer types are described including the notification schedule. Hence, modifications of surveys can be carried out quickly without submitting a new version of the app.

For the ongoing study all participants were asked to provide self-reported ratings of depressive symptoms based on surveys in German language including a digital version of the BDI every two weeks. As for the suicide item of the BDI, the participant is instructed to contact the local Department of Psychiatry if his or her response for this item is 2 or higher (values range from 0-3). In addition, participants were asked to rate their mood and their sleep duration by answering single items on average once a week. For the single mood question (“How is your mood today?”) the participants provide their response via touch-screen on a scale from 1-10 with the anchors 1= “very bad” and 10= “very good”. For the single sleep question (“How many hours did you sleep last night?”) participants provide their response via touch-screen on a scale from 0 to 13 hours. For all self-reported data, the app sends out weekly push notifications on a random basis during daytime with a variance of two days or every two weeks in case of the BDI. Participants are instructed that answering all questions is optional and they are free to choose their time of answering whenever items are made available. No partial events are stored, i.e. if a participant cancels a survey the answers are discarded.

A sleep event is essentially the duration of sleep, a mood event contains the selected mood value and a BDI event encapsulates the entire BDI questionnaire with all 21 selected item responses.

##### Voice Samples

In addition to single items and the BDI, once a week the user is asked to provide a short voice sample. The recording lasts a minimum of one and a maximum of three minutes, but can be interrupted at any time. The participant is instructed to record a free report on his well-being during the last week using the devices microphone, while the app checks the background noise level and simultaneously provides visual feedback on the sufficiency of the sound level of the voice. In this way, a weekly voice sample is obtained while fully preserving the privacy of each individual since each participant may freely decide on what content to report. On iOS we slightly adapted the ResearchKit Audio Task to our needs, on Android we had to redevelop the function, because the SurveyKit Framework lacks this feature. The audio file is recorded in MP4 format, encrypted and sent to the ReMAP server as soon as a wifi connection is available.

#### Passive Sensor-based Data Types

##### Steps and Distances

Information about steps and distance walked are retrieved daily from Apple HealthKit or Google Fit. Internally the corresponding framework uses the smartphone sensors i.e. the pedometer, accelerometer and GPS in case of Android for better distance estimates. A major advantage of this approach is that both event types can be retrieved without draining the battery and data is comparable with data from other studies using the same underlying framework. In detail, one event of steps or distance includes the timestamps for start and end as well as the number of steps or the distance walked in meters in that specific timeframe, meaning the framework detects when the user starts and stops moving and counts that as one event. For these event types access to Apple Health or Google Fit is required as well as access to GPS position for Android only, as mentioned above.

##### Global Positioning System and Accelerometer

GPS location data is collected if a significant location change takes place, this function is provided by the respective platform, so that no permanent background process is necessary. Each location event consists of longitude and latitude coordinates and a timestamp.

For the monitoring of acceleration data, we aimed to implement a technical solution allowing for a meaningful trade-off between sufficient temporal resolution to continuously track the participants physical activity on the one hand and lowest possible battery consumption as well as efficient data storage strategy on the other hand. For iOS the existing cache for acceleration data which is stored for three days on the device is used. On Android the procedure is more complicated since no similar cache for acceleration data exists on Android and a permanent background process is not permitted by the OS. As a compromise ReMAP attempts to retrieve accelerometer sensor data at least every 15 minutes for 3 minutes on Android, in order to not drain the battery too much. Acceleration data is provided as (x, y, z) vector and comes in meters per seconds squared (m/s^2). On iOS the sensor provides the data typically in 60 Hz, on Android it is between 5-100 Hz, depending on the sensor type and the state the phone is currently in. To reduce the amount of incoming data, we collect only one value per second and calculate the average of the absolute values of all values of one second. This provides an indication of whether the phone has been moved or not. For storage and data handling reasons, 4 hours of acceleration data are combined into one event. This should be taken into account when interpreting the acceleration events displayed in this paper.

#### Data Encryption, Security and Privacy

Since the ReMAP app collects highly sensitive personal data, it is important to protect the data from unauthorized access with the highest level of security. For this reason, ReMAP is designed so that data is encrypted at every time. The standard procedure for asynchronous encryption is used, which is also the same as for end-to-end encryption. More precisely, we use a combination of AES-256 and elliptic curves. The public key is provided by the app so that the data can be encrypted on the device. Synchronous keys for encrypting large amounts of data are generated on the device when required. These data encryption keys are then encrypted with the public key, also known as key encryption key, so that the app cannot read the data itself. Only the server can decrypt the data with the corresponding private key. The ReMAP backend runs on university servers and is administered exclusively by the research staff of the study project. The server provides a REST API to upload JSON formatted data using industry standard SSL/TLS encryption for data in transfer. As mentioned above, the server also provides the surveys as JSON files for greater flexibility and updates at runtime.

### Analysis

The user adherence was assessed by means of retention rates and total participation duration. First, the percentage of participants that reached the required minimum participation duration of two weeks was investigated. In a subsequent step, the rate of participants reaching a participation duration of one year (optional to receive second financial compensation) was assessed, as well as the total participation duration in order to further investigate adherence and thus acceptance of the app. Another measure for user acceptance was the rate of transferred active data events. This measure provides insights into the motivation of participants to actively engage in ReMAP (independent from financial compensation). These measures were compared across clinical and non-clinical subgroups in order to investigate a potential sampling bias (e.g. lower motivation for the transfer of active data in subgroups).

The transfer rate of passive data events was used as a measure for technical feasibility of the background functioning of passive data collection. To investigate a potential dependency between active and passive data transfer, we assessed the correlation between the rate of days with passive data events and rate of days with active data events. Measures of technical feasibility were compared between iOS and Android users.

In detail, participation duration was calculated as the time difference between the installation timestamp and the most recent event of a user. The rate of days on which active or passive events were sent were calculated relative to the total duration of participation of a user (days with event divided by total days).

Due to highly skewed distributions in the data, non-parametric tests were applied for all analyses. Spearman rank test was used for correlations, while Mann-Whitney-U and Kruskal-Wallis tests were used for the comparison of groups. All analyses were undertaken in SPSS (IBM, version 26), assuming a significance threshold of p<.05.

## Results

### Adherence

Out of n=1013 total installations a rate of 98.42% provided a valid study ID. Participants with an invalid ID (n=16) were excluded from all further calculations. The remaining n=997 participants had the app installed for an average duration of 150.08 days (SD=128.76).

A rate of 90.25% of the participants sent data for the required minimum time interval of at least two weeks, while 33.09% had the app installed for at least a year. For calculation of these rates all participants were regarded for which installation of the ReMAP app was at least two weeks or one year before data freeze and could thus possibly reach either duration (n=995 and n=275 respectively).

### Amount of Data Collected

#### Active data

An average of 34.59 active events (spanning questionnaires, single-items, and voice samples) were sent by each participant (SD=49.38; range 0-552). The number of unique days on which participants sent active data was on average 13.22 (SD=21.09; range 0-260). Table 1 shows the number of active events across data entities. Participants sent active data on average on 12.60% (SD=18.23%) of their total days of participation.

**Table 1.**
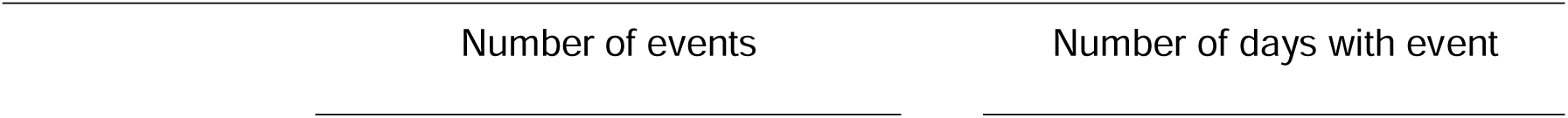
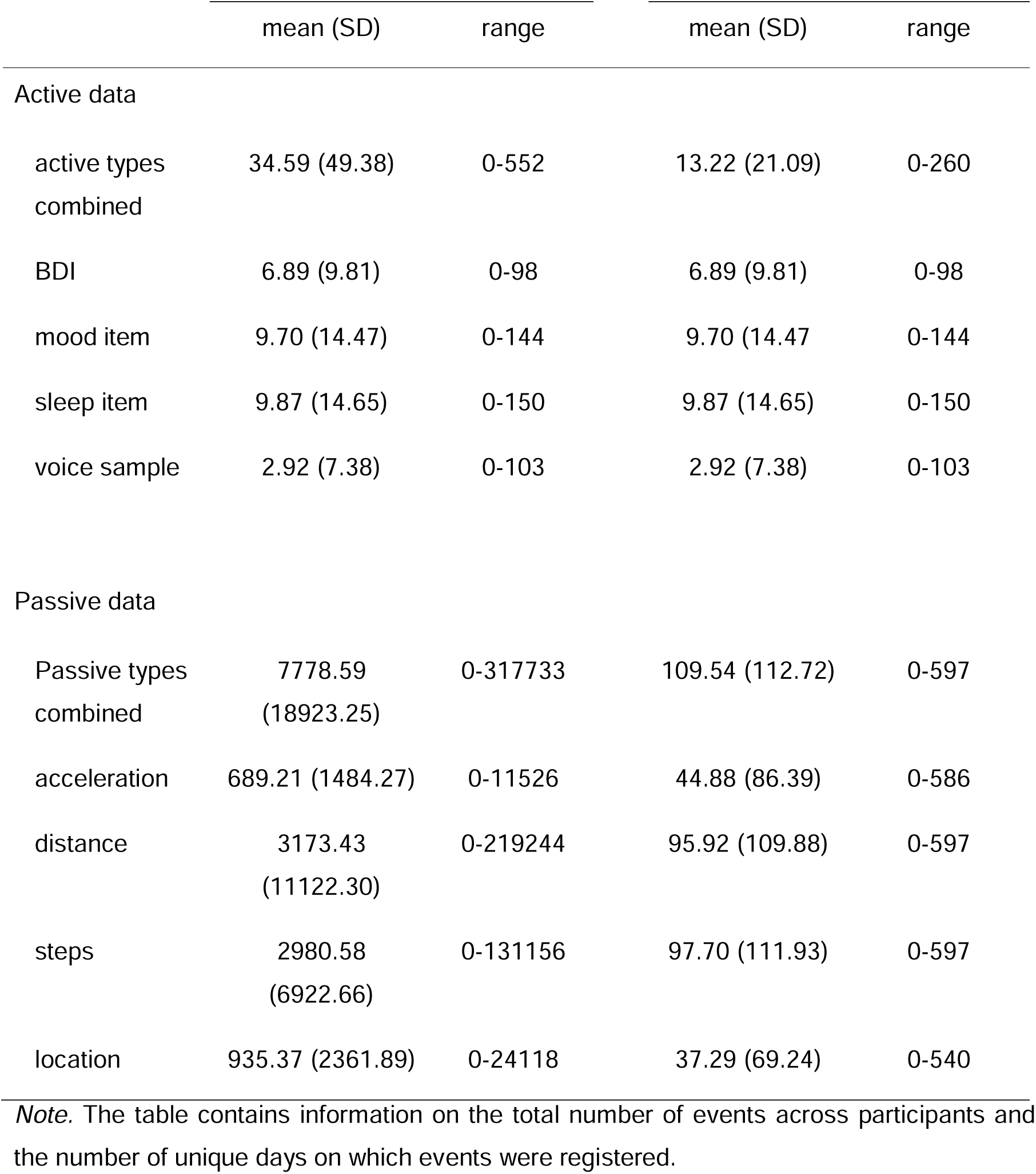
Summary statistics of incoming data events per participant across event types.

#### Passive data

An average of 7778.59 events of passive data were collected for each participant (SD=18923; range 0-317733), resulting in an average of 51.83 passive events per day per participant. Descriptive statistics of registered passive events across data entities is shown in Table 1. The average rate of days on which a passive data event was sent was 73.40% (SD=33.73%) based on the total participation duration.

### Data transfer across clinical and non-clinical subgroups

Diagnosis information (including affirmation of the lack of a clinical diagnosis) was available for n=994 participants that were included in the following analyses. Patient groups included were major depressive disorder (MDD, n=409), bipolar disorder (BD, n=48), anxiety disorder (AD, n=58), and psychotic disorders (PD, n=21). Further, n=458 HC participants were included. HC and different patient subgroups did not differ in regard to total duration of participation (χ^2^(4)=5.654, p=.227) or in regard to the rate of days with active events (χ^2^(4)=3.958, p=.412). Clinical subgroups were found to differ significantly in the rate of days with passive events (χ^2^(4)=11.325, p=.023). However, Bonferroni-corrected post-hoc tests did not reach significance for any pairwise comparisons (all p>.175).

In sum, findings indicate that HC, MDD, BD, AD, and SCZ app users do not differ in regard to their total duration of participation, the rate of active data transfer, or the rate of passive data transfer (Figure 2a).

**Figure 1.**
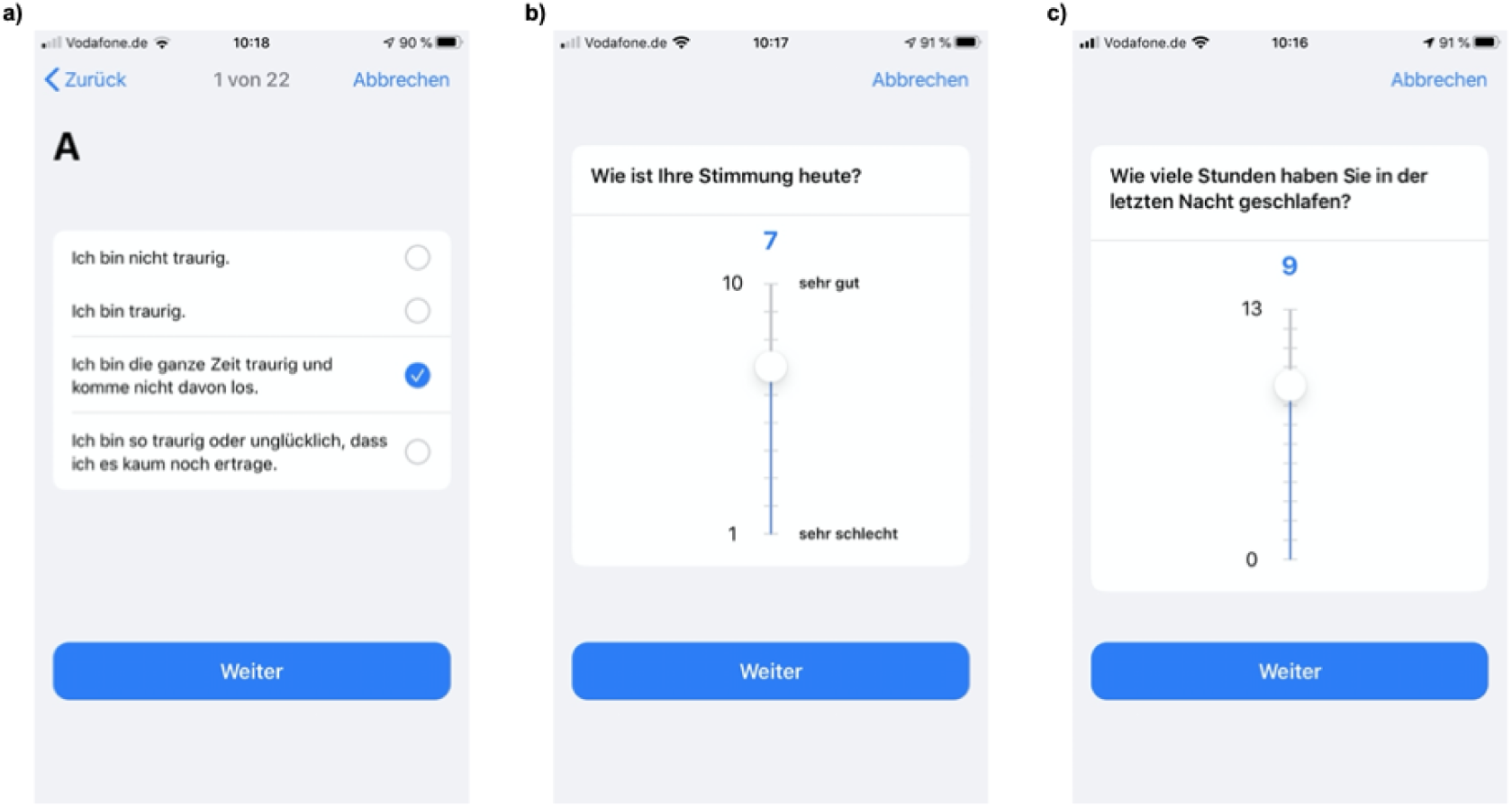
Screenshots of ReMAP surveys are presented as displayed on the participants smartphone to demonstrate the collection routine of actively self-reported data on depressive symptoms (Images recorded on an iPhone 8): a) First item of the mobile version of the BDI questionnaire conducted every two weeks; b) single-item subjective mood rating on a scale from 1-10 conducted on average once a week; c) single-item rating of sleep duration in hours (0-13) conducted once a week. All items are implemented in German language.

**Figure 2.**
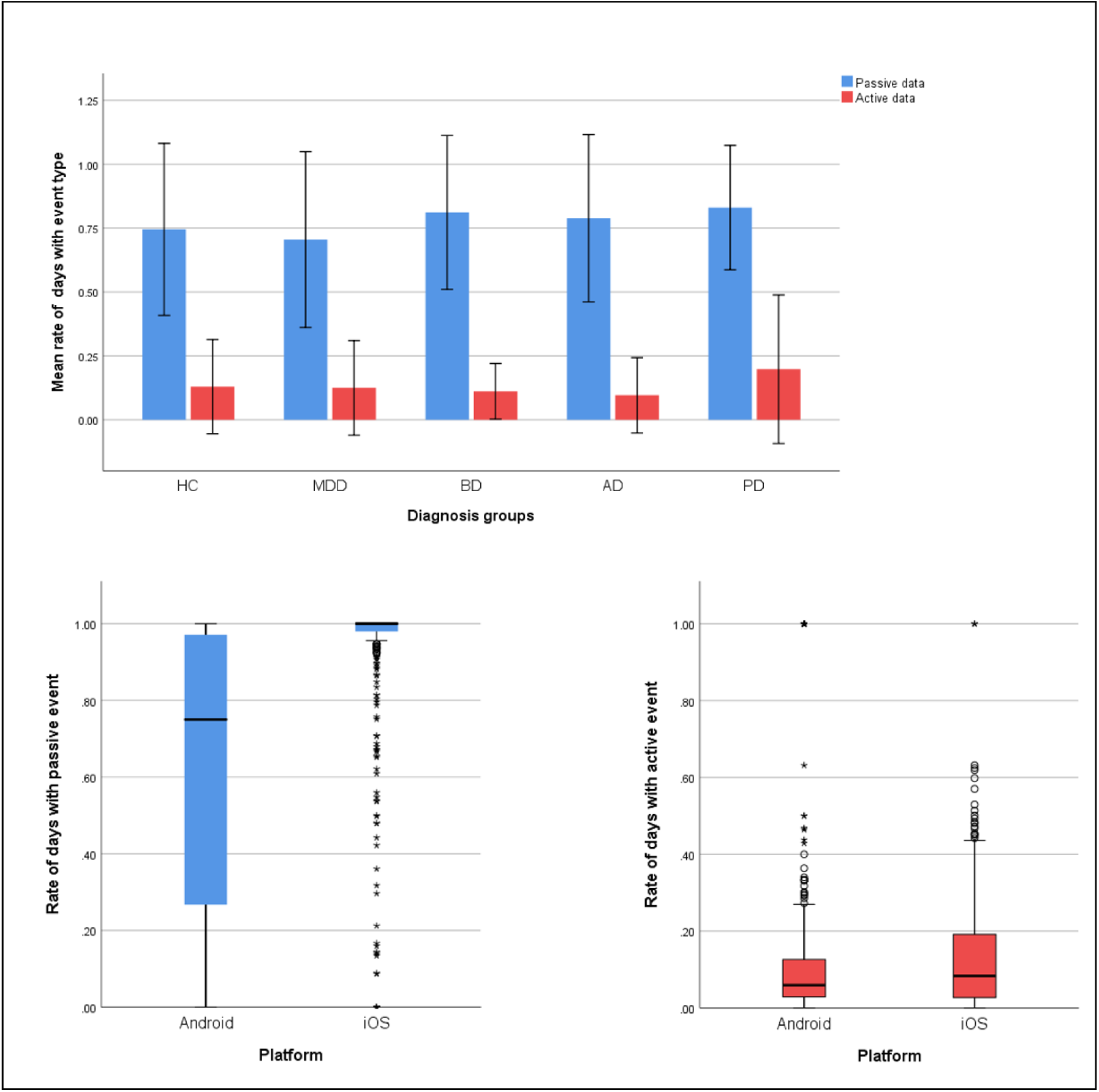
Rate of days with data transfers across participant subgroups and user platforms. Different patient and healthy control subgroups are compared in panel a) in regard to their rate of days with active and passive data transfers. Error bars represent standard deviations. The distribution of this rate of data transfers across user platforms is shown in panel b) for passive events and in panel c) for active event types. HC, healthy control; MDD, major depressive disorder; BD, bipolar disorder; AD, anxiety disorder; PD, psychotic disorder.

### Data transfer across operating systems

Among participants we identified n=632 Android users (63.39%) and n=360 iOS users. Another n=5 users switched from one platform to the other during study participation. These users were excluded from the following analyses.

The mean rate of days with any incoming passive event across all participants was 73.50%. We found this rate to be significantly lower in Android (63.04%) as compared to iOS (91.85%) users (U=46307, p<.001). This pattern of results was consistent when investigating acceleration, distance, steps, and location data separately (all p<.001). The distribution of the rate of days with events across operating systems is shown in Figure 2b for passive event types and in Figure 2c for active event types.

Across all users combined, the rate of days on which participants sent active data correlated positively with the rate of days on which passive data were sent (rho=.165, p<.001). This association was driven by Android users (rho=.221, p<.001), while no significant association was found for iOS users (rho=-.014, p=.798) indicating that specifically in Android users the continuity of passive data transfer was higher the more regularly active data were sent.

## Discussion

The present work provides a detailed description of the development and implementation of the ReMAP app. Evidence is presented that the continuous assessment of activity and mood via active and passive data collection using ReMAP is technically feasible and that participants generally adhere to the app installation and usage.

It is crucial to investigate the technical feasibility of a research app such as ReMAP in order to detect potential technical hazards, such as global performance issues (making the user phone slow or crashing the OS) or noticeable drainage of the battery. Technical feasibility can therefore also be seen as a crucial prerequisite of user acceptance. An important marker for the feasibility is the rate of participants that sent passive data for a duration of at least two weeks, as this was the primary precondition for compensation. The rate of approximately 90% of users participating for at least two weeks and transferring data during this period demonstrates good adherence in general. Furthermore, considering the adherence rate, it appears reasonable to assume acceptable technical feasibility of ReMAP in terms of global performance even though we did not assess participants experience with the app. The remaining 10% that did not reach a two-week duration may in part stem from technical hazards, such as the app not running properly or the change of a phone without notification (Ramsey, Wetherell, Depp, Dixon, & Lenze, 2016). However, another reason could be a late decision to abort study participation due to other motivational reasons that ultimately resulted in deinstallation of the app or withdrawal of essential app permissions. Based on the current data it is impossible to differentiate the proportions of participants ending data transfer due to technical hazards or due to other reasons. However, in previous studies on smartphone-based monitoring in affective disorders technical problems are the most frequently reported reason for adherence discontinuation (Dogan et al., 2017).

Another proxy for technical feasibility is the continuous (as opposed to intermitting) transfer of passive data, while it should be taken into account that a temporarily or permanent modification or withdraw of app permissions by the participant might have biased the observed data transfer rates in the present study. Importantly, we obtained passive events on average on 73.40% of the days of the total participation duration of users. This finding further supports the notion of the ReMAP app being technically feasible in general. However, comparisons of user platforms revealed considerable heterogeneity with regard to the continuity of data transfer between operating systems. While continuity of passive data transfer appeared excellent in iOS users, a considerably lower rate of tracked passive events per day was observed in Android users indicating potential for improvement in the app functioning on Android devices. The finding that specifically for Android users the continuity of passive data was positively associated with the continuity of active data could point to difficulties of background data transfers if the app is not actively opened. Technical challenges of background functioning of digital phenotyping apps are well documented and are further facilitated by rapid progression of operating systems consistently changing software environments (Petter, Hirsch, Mushtaq, Hevesi, & Lukowicz, 2019). Techniques like silent push notifications could be used to improve data collection on Android devices as demonstrated in previous studies (Nishiyama et al., 2020).

Generally, the overall achieved participation durations can be estimated as relatively long, taking into account that financial compensation was humble and considering that low retention rates in app usage are well documented (Bauer et al., 2020). A study investigating smartphone-based symptom self-reports in 126 depressed adults found that approximately 50% of users removed the app within the first two weeks of the study which appears to match the observed reduction in participation rates over time (Wahle, Kowatsch, Fleisch, Rufer, & Weidt, 2016). Further, despite the lack of additional compensation for the transfer of active data, we registered the transfer of a vast amount of single-item answers, questionnaire data, and even a relatively large number of voice samples. This demonstrates that the acceptance/willingness of participants not only to have passive data collected but also to actively engage in the ReMAP app seems to be generally given. Importantly, the rate at which participants provided active data did not differ between healthy controls, affective disorder participants or other psychiatric conditions. This points to comparable acceptance and engagement across clinical and non-clinical subgroups and therefore indicates that the data collection is not subject to a systematic assessment bias (e.g. due to depressive symptomatology).

### Strengths and Limitations

A major strength of the presented study is the comparatively large transdiagnostic sample, including a variety of clinical subgroups. Besides clinical diversity, we demonstrated the general acceptance and technical feasibility of ReMAP across platforms broadly making it available to users with different technical endowment. As recent reviews about smartphone-based monitoring in mental health research demonstrate, previous sample sizes range from 1 to 280 participants (Bardram & Matic, 2020; Bauer et al., 2020; Cornet & Holden, 2018; Dogan et al., 2017), thus making our sample by far the largest. This increases the generalizability of our findings. Further, different active and passive data modalities were combined resulting in an array of data types that can be used for subsequent research questions. In addition the app usage durations of previous studies also vary considerably, ranging from 1 to 365 days (Bardram & Matic, 2020). Both, the number of cases, as well as the number of data types, and the relatively long participation duration in our study are in this combination up to now unprecedented in this field and can markedly facilitate the training of future machine learning algorithms, crucial for predictive modelling.

An important limitation of the current findings is that some qualities were not measured directly but indirect measures were used as proxies. For example, the participation duration was calculated as the difference between the installation timestamp and the most recent event transmitted. However, it is indefinite if users discontinued participation intentionally or if technical difficulties caused the app to stop sending data. Thus, the use of the rate of days with passive events as a measure for continuity of data transfer and thus technical quality is limited as it is dependent on the calculated duration. Therefore, the continuity of data transfer can only be interpreted up to the time point of the last data event transferred. Further, participants were not directly surveyed concerning either their acceptance of the ReMAP app nor regarding the occurrence of technical difficulties or oddities. One possibility for future research would be the application of a specialized usability questionnaire in ReMAP users as done e.g. by Bardram and colleagues in a smartphone study in bipolar patients (Bardram et al., 2013). Another important limitation is that the passive data has not been validated yet.

### Future directions

In a subsequent step the utility of the obtained data for prediction modeling needs to be further investigated (Hays, Farrell, & Torous, 2019). This includes feature engineering particularly of the passive data types but also in regard to the audio samples. Potentially, imputation and interpolation approaches are useful to improve data quality (Mennis, Mason, Coffman, & Henry, 2018). Ultimately, the predictive qualities of the data for clinically relevant changes in affective symptomatology need to be evaluated. The design of the current study (with ReMAP adding on to large existing longitudinal imaging cohort studies) yields the promising possibility to investigate the multimodal prediction of symptom trajectories by combining ReMAP data with genetic and deep phenotyping data (e.g. derived from brain imaging, neuropsychological tests, and clinical ratings). In the long-term these multimodal data may aid to build models utilizable for precision medicine (Robinson, 2012).

### Conclusions

The adherence rates, as well as the high amount and rate/continuity of passive data provided by participants, indicate that the ReMAP app is in principle technically feasible. However, there seems to be room for improvement regarding the background data transfer of passive data on Android devices. Further, long participation durations and a high number of active data transfers could be achieved considering relatively cost-effective compensation, which demonstrates good acceptance of the ReMAP app.

In summary, the ReMAP app is well accepted by users and constitutes a technically feasible and highly cost-efficient tool for the assessment of passive sensor-based activity and self-reported changes in affective symptoms. The feasibility and acceptance was demonstrated across a wide variety of clinical subgroups including affective, anxiety, and psychotic disorders, as well as healthy control participants.

## Data Availability

Data will be made available upon request

## Ethics Statement

All participants received financial compensation. The study was approved by the local institutional review board (IRB = the ethics committee of the Medical Faculty of the University of Münster, Germany) and written informed consent was obtained from all participants before study participation.

## Acknowledgments and Funding

Funding was provided by the DFG – Projectnumber 44541416-TRR 58 (CRC-TRR58, Projects C09 and Z02 to Udo Dannlowski) and the Interdisciplinary Center for Clinical Research (IZKF) of the medical faculty of Münster (Grant Dan3/012/17 to Udo Dannlowski and SEED 11/19 to NO), as well as the “Innovative Medizinische Forschung” (IMF) of the medical faculty of Münster (Grants OP121710 to NO and TH; LE121703 and LE121904 to EJL).

Further DFG funding in the context of the Forschungsgruppe/Research Unit FOR2107 was received by: Tilo Kircher (speaker FOR2107; DFG grant numbers KI 588/14-1, KI 588/14-2, KI 588/15-1, KI 588/17-1), Udo Dannlowski (co-speaker FOR2107; DA 1151/5-1, DA 1151/5-2, DA 1151/6-1), Axel Krug (KR 3822/5-1, KR 3822/7-2), Igor Nenadic (NE 2254/1-2), Carsten Konrad (KO 4291/3-1), Marcella Rietschel (RI 908/11-1, RI 908/11-2), Markus Nöthen (NO 246/10-1, NO 246/10-2), Stephanie Witt (WI 3439/3-1, WI 3439/3-2), Andreas Jansen (JA 1890/7-1, JA 1890/7-2), Tim Hahn (HA 7070/2-2, HA7070/3, HA7070/4), Bertram Müller-Myhsok (MU1315/8-2), Astrid Dempfle (DE 1614/3-1, DE 1614/3-2), Petra Pfefferle (PF 784/1-1, PF 784/1-2), Harald Renz (RE 737/20-1, 737/20-2), Carsten Konrad (KO 4291/4-1).

EJL was supported by the Christiane Nüsslein-Vollhard Foundation.

Principal investigators in the FOR2107 consortium that are not co-authors of the current paper are: Work package (WP) 6, multi-method data analytics: Bertram Müller-Myhsok, Astrid Dempfle. Central project (CP) 1, biobank: Petra Pfefferle, Harald Renz. CP2, administration: Carsten Konrad.

Henrike Bröhl, Bruno Dietsche, Rozbeh Elahi, Jennifer Engelen, Sabine Fischer, Jessica Heinen, Svenja Klingel, Felicitas Meier, Torsten Sauder, Annette Tittmar, Dilara Yüksel (Dept. of Psychiatry, Marburg University). Mechthild Wallnig, Rita Werner (Core-Facility Brainimaging, Marburg University). Carmen Schade-Brittinger, Maik Hahmann (Coordinating Centre for Clinical Trials, Marburg). Michael Putzke (Psychiatric Hospital, Friedberg). Rolf Speier, Lutz Lenhard (Psychiatric Hospital, Haina). Birgit Köhnlein (Psychiatric Practice, Marburg). Peter Wulf, Jürgen Kleebach, Achim Becker (Psychiatric Hospital Hephata, Schwalmstadt-Treysa). Ruth Bär (Care facility Bischoff, Neukirchen). Matthias Müller, Michael Franz, Siegfried Scharmann, Anja Haag, Kristina Spenner, Ulrich Ohlenschläger (Psychiatric Hospital Vitos, Marburg). Matthias Müller, Michael Franz, Bernd Kundermann (Psychiatric Hospital Vitos, Gießen). Katharina Förster, Kordula Vorspohl, Bettina Walden, Dario Zaremba, Lena Waltemate, Dominik Grotegerd, Joscha Böhnlein, Tiana Borgers, Verena Enneking, Stella Fingas, Marius Gruber, Carina Hülsmann, Hannah Lemke, Susanne Meinert, Maike Richter, Lisa Sindermann, Katharina Thiel (Dept. of Psychiatry, University of Münster). Harald Kugel, Walter Heindel, Birgit Vahrenkamp (Dept. of Clinical Radiology, University of Münster). Gereon Heuft, Gudrun Schneider (Dept. of Psychosomatics and Psychotherapy, University of Münster). Thomas Reker (LWL-Hospital Münster). Gisela Bartling (IPP Münster). Ulrike Buhlmann (Dept. of Clinical Psychology, University of Münster).

Helene Dukal, Christine Hohmeyer, Lennard Stütz, Viola Schwerdt, Fabian Streit, Josef Frank, Lea Sirignano (Dept. of Genetic Epidemiology, Central Institute of Mental Health, Medical Faculty Mannheim, Heidelberg University). Stefanie Heilmann-Heimbach, Stefan Herms, Per Hoffmann (Institute of Human Genetics, University of Bonn, School of Medicine & University Hospital Bonn). Andreas J. Forstner (Institute of Human Genetics, University of Bonn, School of Medicine & University Hospital Bonn; Centre for Human Genetics, Marburg University).

Anastasia Benedyk, Miriam Bopp, Roman Keßler, Maximilian Lückel, Verena Schuster, Christoph Vogelbacher (Dept. of Psychiatry, Marburg University). Jens Sommer, (Core-Facility Brainimaging, Marburg University). Thomas W.D. Möbius (Institute of Medical Informatics and Statistics, Kiel University).

Julian Glandorf, Fabian Kormann, Arif Alkan, Fatana Wedi, Lea Henning, Alena Renker, Karina Schneider, Elisabeth Folwarczny, Dana Stenzel, Kai Wenk, Felix Picard, Alexandra Fischer, Sandra Blumenau, Beate Kleb, Doris Finholdt, Elisabeth Kinder, Tamara Wüst, Elvira Przypadlo, Corinna Brehm (Comprehensive Biomaterial Bank Marburg, Marburg University).

We are further deeply indebted to all participants of this study.

## References

Bardram, J. E. (2020). The CARP Mobile Sensing Framework-A Cross-platform, Reactive, Programming Framework and Runtime Environment for Digital Phenotyping.

Bardram, J. E., Frost, M., Szántó, K., Faurholt-Jepsen, M., Vinberg, M., & Kessing, L. V. (2013). Designing mobile health technology for bipolar disorder: A field trial of the MONARCA system. Conference on Human Factors in Computing Systems - Proceedings, 2627–2636. https://doi.org/10.1145/2470654.2481364

Bardram, J. E., & Matic, A. (2020). A Decade of Ubiquitous Computing Research in Mental Health. IEEE Pervasive Computing, 19(1), 62–72. https://doi.org/10.1109/MPRV.2019.2925338

Bauer, M., Glenn, T., Geddes, J., Gitlin, M., Grof, P., Kessing, L. V., … Whybrow, P. C. (2020). Smartphones in mental health: a critical review of background issues, current status and future concerns. International Journal of Bipolar Disorders, 8(2), 1–19. https://doi.org/10.1186/s40345-019-0164-x

Beck, A. T., Ward, C. H., Mendelson, M., Mock, J., & Erbaugh, J. (1961). An Inventory for Measuring Depression. Archives of General Psychiatry, 4, 561–571. https://doi.org/10.1001/archpsyc.1961.01710120031004

Cornet, V. P., & Holden, R. J. (2018). Systematic review of smartphone-based passive sensing for health and wellbeing. Journal of Biomedical Informatics, 77, 120–132. https://doi.org/10.1016/j.jbi.2017.12.008

Coyne, J. C., Thompson, R., & Racioppo, M. W. (2001). Validity and efficiency of screening for history of depression by self-report. Psychological Assessment, 13(2), 163–170.

Dogan, E., Sander, C., Wagner, X., Hegerl, U., & Kohls, E. (2017). Smartphone-Based Monitoring of Objective and Subjective Data in Affective Disorders: Where Are We and Where Are We Going? Systematic Review. Journal of Medical Internet Research, 19(7), e262. https://doi.org/10.2196/jmir.7006

Faurholt-Jepsen, M., Brage, S., Vinberg, M., Christensen, E. M., Knorr, U., Jensen, H. M., & Kessing, L. V. (2012). Differences in psychomotor activity in patients suffering from unipolar and bipolar affective disorder in the remitted or mild/moderate depressive state. Journal of Affective Disorders, 141(2–3), 457–463. https://doi.org/10.1016/j.jad.2012.02.020

Faurholt-Jepsen, M., Vinberg, M., Frost, M., Christensen, E. M., Bardram, J. E., & Kessing, L. V. (2015). Smartphone data as an electronic biomarker of illness activity in bipolar disorder. Bipolar Disorders, 17(7), 715–728. https://doi.org/10.1111/bdi.12332

Goltermann, J., Emden, D., Leehr, E. J., Dohm, K., Redlich, R., Dannlowski, U., … Opel, N. (2020). Validation of smartphone-based assessments of depressive symptoms using the Remote Monitoring Application in Psychiatry (ReMAP). MedRxiv, 2020.08.27.20183418. https://doi.org/10.1101/2020.08.27.20183418

Hays, R., Farrell, H. M., & Torous, J. (2019). Mobile apps and mental health: Using technology to quantify real-time clinical risk. Current Psychiatry, 18(6), 37–41.

Kleiman, E. M., Turner, B. J., Fedor, S., Beale, E. E., Picard, R. W., Huffman, J. C., & Nock, M. K. (2018). Digital phenotyping of suicidal thoughts. Depression and Anxiety, 35(7), 601–608. https://doi.org/10.1002/da.22730

Mennis, J., Mason, M., Coffman, D. L., & Henry, K. (2018). Geographic imputation of missing activity space data from ecological momentary assessment (EMA) GPS positions. International Journal of Environmental Research and Public Health, 15(12). https://doi.org/10.3390/ijerph15122740

Nishiyama, Y., Ferreira, D., Sasaki, W., Okoshi, T., Nakazawa, J., Dey, A. K., & Sezaki, K. (2020). Using iOS for inconspicuous data collection: A real-world assessment. Adjunct - Proceedings of the 2020 ACM International Joint Conference on Pervasive and Ubiquitous Computing and Proceedings of the 2020 ACM International Symposium on Wearable Computers, 261–266. https://doi.org/10.1145/3410530.3414369

Onnela, J.-P., & Rauch, S. L. (2016). Harnessing Smartphone-Based Digital Phenotyping to Enhance Behavioral and Mental Health. Neuropsychopharmacology, 41, 1691–1696. https://doi.org/10.1038/npp.2016.7

Petter, O., Hirsch, M., Mushtaq, E., Hevesi, P., & Lukowicz, P. (2019). Crowdsensing under recent mobile platform background service restrictions - A practical approach.

UbiComp/ISWC 2019- - Adjunct Proceedings of the 2019 ACM International Joint Conference on Pervasive and Ubiquitous Computing and Proceedings of the 2019 ACM International Symposium on Wearable Computers, 793–797. https://doi.org/10.1145/3341162.3344867

Ramsey, A. T., Wetherell, J. L., Depp, C., Dixon, D., & Lenze, E. (2016). Feasibility and Acceptability of Smartphone Assessment in Older Adults with Cognitive and Emotional Difficulties. Journal of Technology in Human Services, 34(2), 209–223. https://doi.org/10.1080/15228835.2016.1170649

Robinson, P. N. (2012). Deep phenotyping for precision medicine. Human Mutation, 33, 777– 780. https://doi.org/10.1002/humu.22080

Russell, M. A., & Gajos, J. M. (2020). Annual Research Review: Ecological momentary assessment studies in child psychology and psychiatry. Journal of Child Psychology and Psychiatry, 61(3), 376–394. https://doi.org/10.1111/jcpp.13204

Torous, J., Choudhury, T., Barnett, I., Keshavan, M., & Kane, J. (2020). Smartphone relapse prediction in serious mental illness: a pathway towards personalized preventive care. World Psychiatry, 19(3), 307–308. https://doi.org/10.1002/wps.20798

Torous, J., Larsen, M. E., Depp, C., Cosco, T. D., Barnett, I., Nock, M. K., & Firth, J. (2018). Smartphones, Sensors, and Machine Learning to Advance Real-Time Prediction and Interventions for Suicide Prevention: a Review of Current Progress and Next Steps. Current Psychiatry Reports, 20(51).

Wahle, F., Kowatsch, T., Fleisch, E., Rufer, M., & Weidt, S. (2016). Mobile Sensing and Support for People With Depression: A Pilot Trial in the Wild. JMIR MHealth and UHealth, 4(3), e111. https://doi.org/10.2196/mhealth.5960

