## Supplementary Material for "Technical feasibility and acceptance of the Remote Monitoring Application in Psychiatry (ReMAP)"

*Description of sample cohorts*

As previously described (1) the sample for the current analyses were drawn from several ongoing longitudinal cohorts. Included samples stem from the Marburg/MünsterAffectiveDisorderCohortStudy (MACS, n=584) (2, 3), the MünsterNeuroimageCohort (MNC, n=157) (4, 5), two subsamples of the SFB-TRR58 cohort (total of n=144; Z02 Münster cohort and SpiderVR Münster cohort (6)), the TIP (n=56) cohort, and the SEED 11/19 cohort (n=56) (7). All cohorts comprise healthy control (HC) participants, as well as different patient groups. Both, the MACS and the MNC cohorts include major depressive disorder (MDD) and bipolar disorder (BD) patients under current or former inpatient treatment. The MACS cohort additionally includes psychotic disorder (PD) patients. The SFB-TRR58 sample includes patients with a spider phobia (SP). The TIP sample includes patients with a social anxiety disorder (SAD), MDD, or with comorbid SAD and MDD. The SEED 11/19 cohort includes patients with MDD, BD, and PD that are under current inpatient treatment during recruitment.

### References


1. Goltermann J, Emden D, Leehr EJ, et al.: Validation of smartphone-based assessments of depressive symptoms using the Remote Monitoring Application in Psychiatry (ReMAP). medRxiv 2020; 2020.08.27.20183418[cited 2020 Sep 21] Available from: https://www.medrxiv.org/content/10.1101/2020.08.27.20183418v1

2. Vogelbacher C, Möbius TWD, Sommer J, et al.: The Marburg-Münster Affective Disorders Cohort Study (MACS): A quality assurance protocol for MR neuroimaging data. Neuroimage 2018; 172:450–460

3. Kircher T, Wöhr M, Nenadic I, et al.: Neurobiology of the major psychoses: a translational perspective on brain structure and function-the FOR2107 consortium. Eur Arch Psychiatry Clin Neurosci 2018;

4. Opel N, Redlich R, Dohm K, et al.: Mediation of the influence of childhood maltreatment on depression relapse by cortical structure: a 2-year longitudinal observational study. The Lancet Psychiatry 2019; 6:318–326 Available from:

5. Dannlowski U, Kugel H, Grotegerd D, et al.: Disadvantage of Social Sensitivity: Interaction of Oxytocin Receptor Genotype and Child Maltreatment on Brain Structure. Biol Psychiatry 2016; 80:398–405

6. Schwarzmeier H, Leehr EJ, Böhnlein J, et al.: Theranostic markers for personalized therapy of spider phobia: Methods of a bicentric external cross-validation machine learning approach. Int J Methods Psychiatr Res 2020; 29:e1812

7. Richter M, Storck M, Blitz R, et al.: Continuous digital collection of patient-reported outcomes during inpatient treatment for affective disorders - implementation and feasibility. medRxiv 2020; 2020.08.27.20183400
